# Assessing equitable use of large language models for clinical decision support in real-world settings: fine-tuning and internal-external validation using electronic health records from South Asia

**DOI:** 10.1101/2024.06.05.24308365

**Authors:** Seyed Alireza Hasheminasab, Faisal Jamil, Muhammad Usman Afzal, Ali Haider Khan, Sehrish Ilyas, Ali Noor, Salma Abbas, Hajira Nisar Cheema, Muhammad Usman Shabbir, Iqra Hameed, Maleeha Ayub, Hamayal Masood, Amina Jafar, Amir Mukhtar Khan, Muhammad Abid Nazir, Muhammad Asaad Jamil, Faisal Sultan, Sara Khalid

**Affiliations:** Centre for Statistics in Medicine (CSM), Nuffield Department of Orthopaedics, Rheumatology and Musculoskeletal Sciences (NDORMS), University of Oxford, Oxford, United Kingdom; Shaukat Khanum Memorial Cancer Hospital and Research Centre, Lahore, Punjab, Pakistan

**Author notes:** **Corresponding Author:** Sara Khalid, Centre for Statistics in Medicine, Botnar Institute for Musculoskeletal Sciences, Windmill Road, Oxford, OX3 7LD, United Kingdom.

## Abstract

**Objective:** Fair and safe Large Language Models (LLMs) hold the potential for clinical task-shifting which, if done reliably, can benefit over-burdened healthcare systems, particularly for resource-limited settings and traditionally overlooked populations. However, this powerful technology remains largely understudied in real-world contexts, particularly in the global South. This study aims to assess if openly available LLMs can be used equitably and reliably for processing medical notes in real-world settings in South Asia.

**Methods:** We used publicly available medical LLMs to parse clinical notes from a large electronic health records (EHR) database in Pakistan. ChatGPT, GatorTron, BioMegatron, BioBert and ClinicalBERT were tested for bias when applied to these data, after fine-tuning them to a) publicly available clinical datasets I2B2 and N2C2 for medical concept extraction (MCE) and emrQA for medical question answering (MQA), and b) the local EHR dataset. For MCE models were applied to clinical notes with 3-label and 9-label formats and for MQA were applied to medical questions. Internal and external validation performance was measured for a) and b) using F1, precision, recall, and accuracy for MCE and BLEU and ROUGE-L for MQA.

**Results:** LLMs not fine-tuned to the local EHR dataset performed poorly, suggesting bias, when externally validated on it. Fine-tuning the LLMs to the local EHR data improved model performance. Specifically, the 3-label precision, recall, F1 score, and accuracy for the dataset improved by 21-31%, 11-21%, 16-27%, and 6-10% amongst GatorTron, BioMegatron, BioBert and ClinicalBERT. As an exception, ChatGPT performed better on the local EHR dataset by 10% for precision and 13% for each of recall, F1 score, and accuracy. 9-label performance trends were similar.

**Conclusions:** Publicly available LLMs, predominantly trained in global north settings, were found to be biased when used in a real-world clinical setting. Fine-tuning them to local data and clinical contexts can help improve their reliable and equitable use in resource-limited settings. Close collaboration between clinical and technical experts can ensure responsible and unbiased powerful tech accessible to resource-limited, overburdened settings used in ways that are safe, fair, and beneficial for all.

## Introduction

Medical Large Language Models (LLMs) propose to leverage the power of multi-billion-parameter neural networks to unlock, summarise, and present medical information quickly and easily, to boost clinical decision-making. Ultimately, by extracting insights from massive volumes of clinical notes with unprecedented speed and accuracy, LLMs can potentially feed a variety of task-shifting applications including, and not limited to, frontline worker decision-support, clinical trial selection for life-saving treatments, culturally appropriate medical training, medical data discovery and evidence generation.[1–4] If done reliably and speedily, such use of LLMs can benefit over-burdened healthcare systems everywhere, but particularly in resource-limited settings such as South Asia, home to a quarter of the world’s population, and where rural and urban health facilities are largely over-subscribed and under pressure.

However, the technology is still in its infancy and lacks clinical uptake.[5–10] Little is known about how LLMs perform in real-world settings, and if they are fair, safe, ethical, and trusted in bespoke settings.[11] Despite the recent surge in language-based models such as ChatGPT-4,[12] the validation of their clinical applications and robust regulatory debate over their use is still lacking, hindering their uptake within resource-limited healthcare settings.

Electronic health records (EHRs), including clinical notes, represent enormous repositories of information to aid patient care. However, the efficient use of these data sources is impeded by a lack of syntactic, structural, and semantic interoperability and standardization.

LLMs stand ready as invaluable tools to overcome these challenges, promising enhanced interpretation and knowledge retrieval within the intricate landscape of healthcare data.

Often, key subjective information including family history, drug adverse events and social, behavioural, and environmental determinants of health – all of which are commonly required in time-critical decision-making – is well-documented only within the full-text patient notes of EHRs.[13] Used in combination with structured data, this information can provide key contextual, socio-demographic and cultural nuances to improve health care, especially for traditionally marginalised communities with limited health access and representation in health data.

LLMs hold promise to improve healthcare and reduce health disparities through their ability to process these data-rich sources and provide critical information to clinicians. The use of these models could hold particular benefits for traditionally overlooked groups, including women, children, and socio-economically or otherwise deprived populations; however, these impacts are only possible if the underlying models prevent the exacerbation of biases.

Although AI bias can be multi-faceted, two main sources are a) unrepresentative training and testing data, and b) algorithmic bias. Most AI tools have been developed in global North settings using data from high-income demographics.[14] Ensuing models may therefore be under-representative of low-income geographies and populations, resulting in algorithmic assumptions about gender, race, and geography, socio-economic status, etc. Concealed biases within LLMs could have severe repercussions on patient outcomes, and render them unsuitable for use with diverse, global populations.[15–17]

In this paper, we studied the strengths and limitations of LLMs in a real-world, global South clinical setting. We tested, and independently validated, publicly available LLMs on a large local EHR database from South Asia. To assess bias, we compared model performance with and without fine-tuning the model to the local dataset. We assessed both internal and external validation by testing the performance of models fine-tuned on the local EHR dataset on open datasets, and vice versa (Figure 1). Models were used to parse clinical notes. This approach is disease-agnostic and generalisable to other downstream uses of LLMs such as summarisation, inference and more. Key contributions of this study include a demonstration of the challenges and opportunities in the use of LLMs in real-world, resource-limited settings. This work opens up avenues to further study the potential of LLMs to empower clinical decision-making and enable task-shifting which is beneficial for all.

**Figure 1:** Study Design for assessing bias of LLMs in real-world clinical settings. Internal and external validation was undertaken with and without fine-tuning on real-world data.

## Methods

### Data source

The Shaukat Khanum Memorial Cancer Hospital and Research Centre (SKMCH&RC) (www.shaukatkhanum.org.pk) is a secondary and tertiary care hospital network spanning 70 cities in Pakistan. Its electronic health records database contains free-text notes and structured data for 8.2 million actively registered patients (51% women).[18] It is linked with the Punjab Cancer Registry and contains anonymised, de-identified patient-level data on socio-demographics, laboratory results, clinical history, diagnoses, outcomes, prescriptions/dispensations, hospital in-patient procedures, and mortality from December 1994 to the present (1st June 2022). The SKMCH&RC dataset contains two types of free-text notes: DS notes and SOAP notes.

### Inpatient Discharge Summary (DS) Notes

DS notes represent a comprehensive summary of a patient’s hospitalization, including diagnostic information, procedures performed, medications administered, and post-discharge instructions. Patient demographics, admission and discharge dates, primary consultants, and detailed information about the patient’s condition are documented under “Diagnosis During This Admission,” “Background Medical Problem(s),” and “Management During Admission” headings. These data can provide subjective information not necessarily captured in structured codes.

### Subjective, Objective, Assessment, and Plan Notes (SOAP)

SOAP notes offer a structured approach to documenting patient information in 4 sections.

1) Subjective: Patient symptoms, history, and any information provided by the patient or caregiver.
2) Objective: Objective observations, laboratory results, and imaging data.
3) Assessment: Diagnosis, problem list, and a summary of the patient’s health status.
4) Plan: Detailed plans for treatment, medications, follow-up, and any other relevant actions.

SOAP notes provide a nuanced understanding of patient cases, ranging from diagnostic workups to treatment plans. Key entities in the dataset include patient demographics, medical history, diagnostic findings, treatment plans, and follow-up instructions.

### Labelling Clinical Notes

A team of six clinical experts including resident doctors labelled the DS and SOAP notes both for concept extraction and question answering tasks. Using a consensus approach, a label/answer with the highest level of agreement between the team was considered the “true” label. The labelled dataset was double-checked by the resident supervisory doctor. A token was considered to be the smallest unit of text that a given model can read such as a word, sub-word, or character. For concept extraction, each token was labelled using the Inside-Outside-Beginning (BIO) format. For each expression, the first token was labelled with “B” followed by tokens within the expression labelled with “I” and tokens outside the expression labelled with “O”.

### Medical LLMs

In addition to OpenAI’s ChatGPT as a trained general-purpose language model, 4 publicly available medical large language models (Table 1) designed for parsing medical notes were used, namely GatorTron(base), BioMegatron, ClinicalBERT, and BioBERT.[19–22] These open-source models are available pre-trained on extensive medical datasets, allowing them to acquire a nuanced understanding of both medical terminology and English text structures. Pre-training equips the models with a comprehensive grasp of medical concepts and proficiency in medical context and vocabulary. Additional fine-tuning layers allow these models to be used for tasks such as clinical concept extraction, medical relation extraction, natural language inference, semantic textual similarity, medical event prediction, and question answering.

**Table 1:**
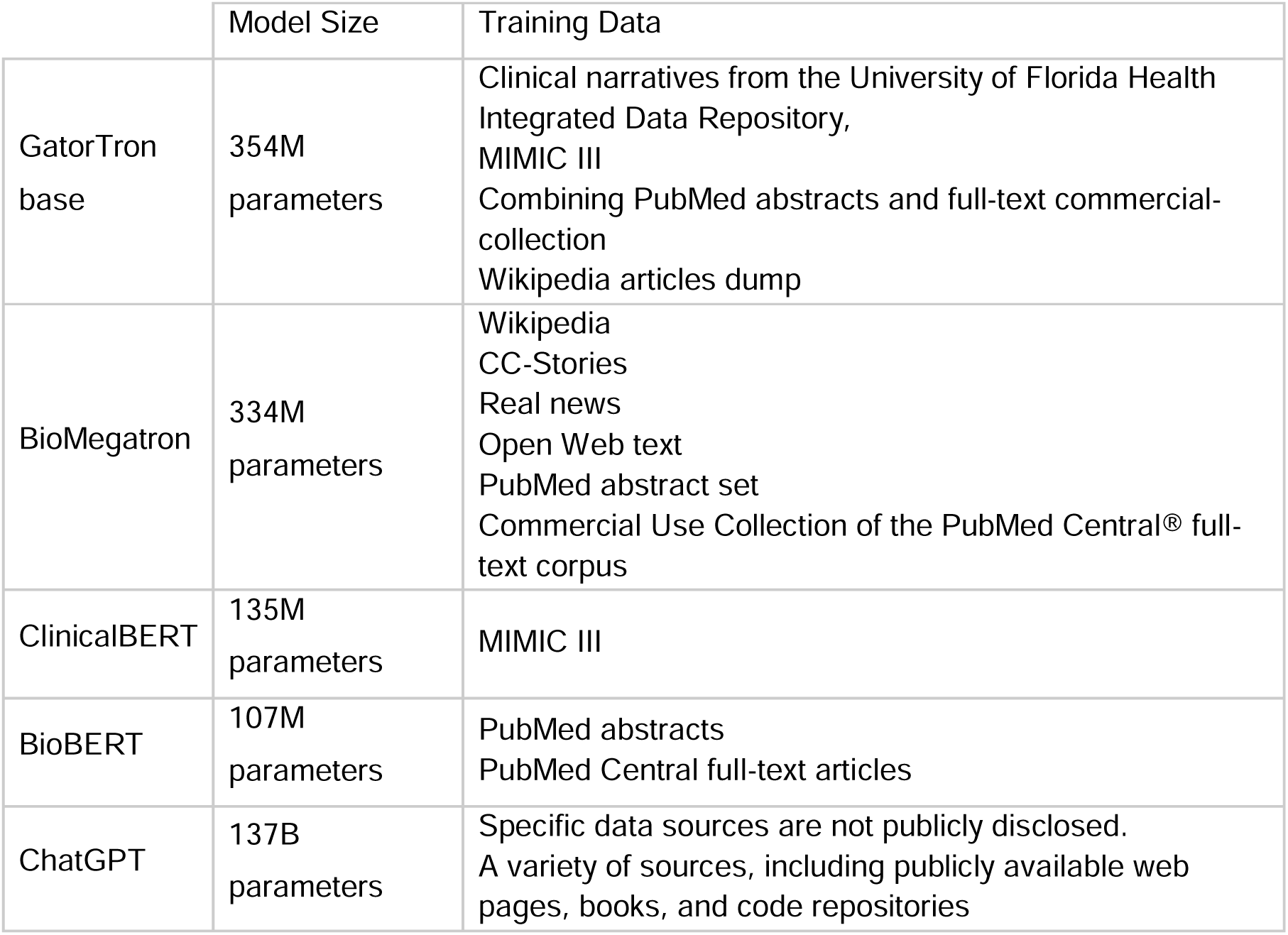
Pre-trained clinical LLMs used in the study.

### Fine-tuning LLMs

Each LLM is available pre-trained on large volumes of data (Table 1). In this study, we fine-tuned each pre-trained LLM for the task of parsing DS and SOAP notes through named entity recognition (NER), which involves identifying specific entities in medical records. Aside from ChatGPT which was trained by OpenAI, each LLM was separately fine-tuned on a) publicly available datasets i2b2 2010 (3 labels: treatment, test, problem), and n2c2 (9 labels: Duration, Frequency, Strength, Form, Route, Dosage, Reason, ADE, Drug), and emrQA (for question answering task), and b) the SKMCH&RC dataset for each corresponding task. Publicly available datasets were pre-segmented into training and testing sets. The SKMCH&RC dataset was randomly split into training (80%) and test (20%) sets.

### Model Performance: Internal and external validation

LLMs fine-tuned on i2b2 2010, n2c2, and emrQA training sets were internally validated on their respective test sets. They were then externally validated on the SKMCH&RC test set. Similarly, LLMs fine-tuned on the SKMCH&RC train set were internally validated on the SKMCH&RC test set and externally validated on the i2b2 2010, n2c2, and emrQA test sets. ChatGPT was tested on the SKMCH&RC dataset without fine-tuning.

### Evaluation of Bias

Model performance was measured using F1 Score, precision, recall, and accuracy for the medical concept extraction and BLEU and ROUGE-L for question answering task. Confusion matrices were produced to assess label-specific misclassification.

All the presented open-source LLMs are accessible through the Hugging Face website at no cost, except for ChatGPT, which requires a paid subscription. To conduct fine-tuning and evaluate model performance, we utilized a Google Colaboratory account with a paid subscription, equipped with an A100 GPU.

## Results

A total of 200 free-text notes, including 50 DS and 150 SOAP notes were randomly extracted from the SKMCH&RC dataset for a two-year period from 01-Jan-2020 to 21-Nov-2021. One note per patient was extracted; the notes represented a patient population including 46% men and 54% women; 89% were adults (defined as aged 19-87 years).

This included 284,445 expert-labelled tokens in BIO format, including 140,841 tokens representing the 3 classes and 143,604 tokens representing the 9 classes. In comparison, the public datasets contained a total number of 2,485,556 BIO tokens, including 1,314,036 tokens representing the 3 classes and 1,171,520 tokens representing the 9 classes. Table 2 displays fine-tuning and internal/external testing dataset statistics.

**Table 2:**
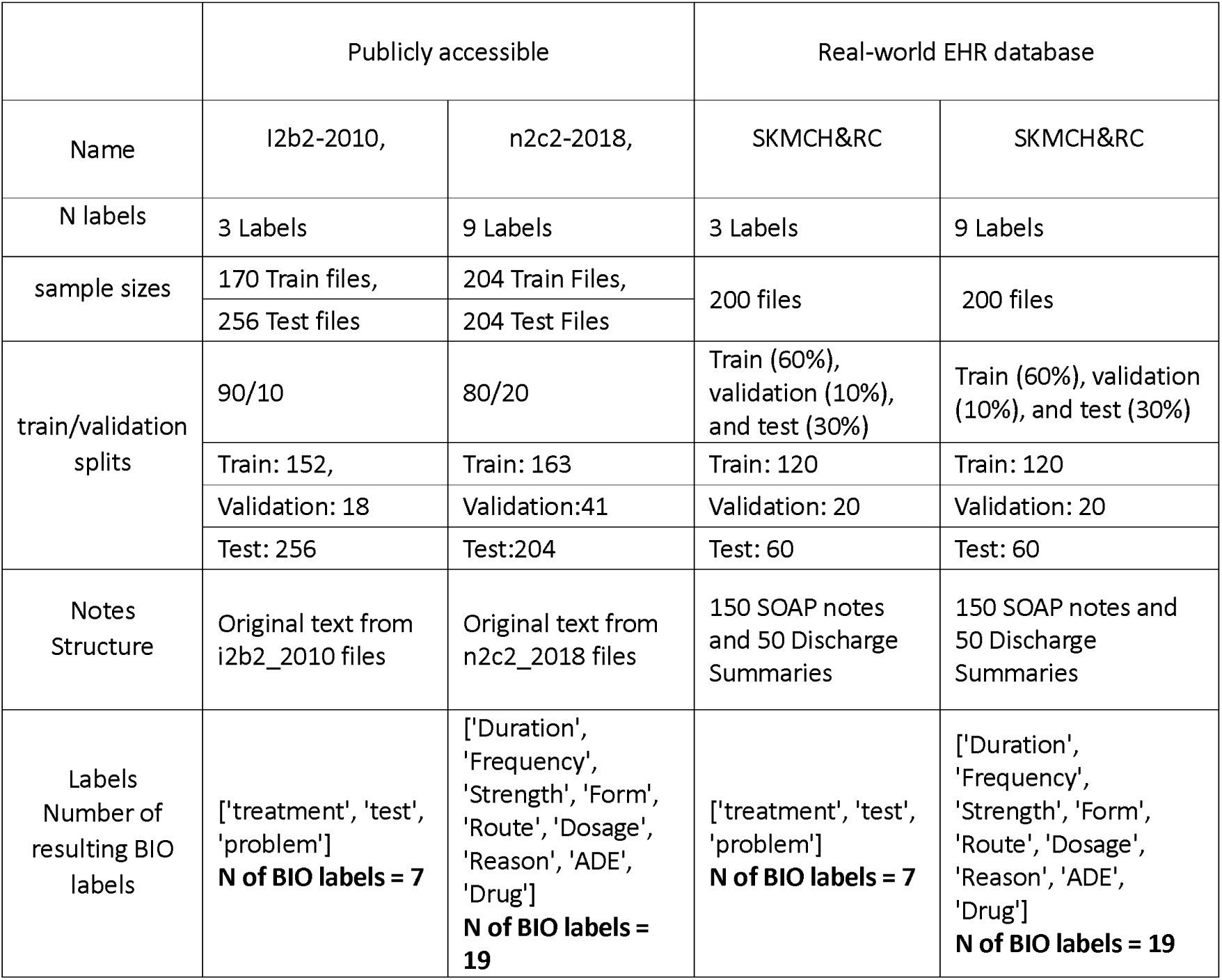
Datasets used for fin-tuning and internal and external validation.

|  | Publicly accessible |  | Real-world EHR database |  |
| --- | --- | --- | --- | --- |
| Name | i2b2-2010, | n2c2-2018, | SKMCH&RC | SKMCH&RC |
| N labels | 3 Labels | 9 Labels | 3 Labels | 9 Labels |
| sample sizes | 170 Train files, | 204 Train Files, | 200 files | 200 files |
|  | 256 Test files | 204 Test Files |  |  |
| train/validation splits | 90/10 | 80/20 | Train (60%), validation (10%), and test (30%) | Train (60%), validation (10%), and test (30%) |
|  | Train: 152, | Train: 163 | Train: 120 | Train: 120 |
|  | Validation: 18 | Validation:41 | Validation: 20 | Validation: 20 |
|  | Test: 256 | Test:204 | Test: 60 | Test: 60 |
| Notes Structure | Original text from i2b2_2010 files | Original text from n2c2_2018 files | 150 SOAP notes and 50 Discharge Summaries | 150 SOAP notes and 50 Discharge Summaries |
| Labels<br>Number of resulting BIO labels | ['treatment', 'test', 'problem']<br>**N of BIO labels = 7** | ['Duration', 'Frequency', 'Strength', 'Form', 'Route', 'Dosage', 'Reason', 'ADE', 'Drug']<br>**N of BIO labels = 19** | ['treatment', 'test', 'problem']<br>**N of BIO labels = 7** | ['Duration', 'Frequency', 'Strength', 'Form', 'Route', 'Dosage', 'Reason', 'ADE', 'Drug']<br>**N of BIO labels = 19** |

### Model Performance

Model performance for internal and external validation is summarised in Table 3.

**Table 3:** Concept extraction model performance for internal and external validation of LLMs fine-tuned on publicly available and SKMCH&RC datasets. In this table, each row indicates whether fine-tuning and testing were conducted on public or SKMCH&RC datasets. By referencing the number of labels in each column, the dataset used can be inferred. For instance, the combination of “public” and “3-label” implies the utilization of the public dataset with a 3-label or 2B2 dataset.

| Model Name | GatorTron<br>Base | BioMegatron | Clinical_BERT | Bio_BERT | ChatGPT | GatorTron_Base | BioMegatron | ClinicalBERT | BioBERT | ChatGPT |
| --- | --- | --- | --- | --- | --- | --- | --- | --- | --- | --- |
| Label Format | 3-Label |  |  |  |  | 9-Label |  |  |  |  |
| Precision<br>Finetuned on Public<br>Tested on Public | **0.8303**<br>[0.8256,<br>0.8368] | **0.8575**<br>[0.8509,<br>0.8619] | **0.8612**<br>[0.8567,<br>0.8651] | **0.8641**<br>[0.8584,<br>0.8704] | **0.8114** | **0.9555**<br>[0.9516, 0.9599] | **0.946**<br>[0.9411,<br>0.9499] | **0.9316**<br>[0.9254,<br>0.9376] | **0.9476**<br>[0.9436,<br>0.9523] | **0.9124** |
| Recall<br>Finetuned on Public<br>Tested on Public | **0.8448**<br>[0.8305,<br>0.8579] | **0.8736**<br>[0.8617,<br>0.8826] | **0.8791**<br>[0.8679, 0.888] | **0.869**<br>[0.8578,<br>0.8788] | **0.76** | **0.9625**<br>[0.9591, 0.9668] | **0.956**<br>[0.9502,<br>0.9605] | **0.9462**<br>[0.9419,<br>0.9512] | **0.9678**<br>[0.9653,<br>0.9714] | **0.6978** |
| F1<br>Finetuned on Public<br>Tested on Public | **0.8375**<br>[0.8288,<br>0.8447] | **0.8655**<br>[0.8569,<br>0.872] | **0.8701**<br>[0.8623,<br>0.8746] | **0.8665**<br>[0.8581,<br>0.8746] | **0.7667** | **0.959**<br>[0.9556, 0.9628] | **0.951**<br>[0.9546,<br>0.9546] | **0.9388**<br>[0.9339,<br>0.9441] | **0.9576**<br>[0.9548,<br>0.9615] | **0.7496** |
| Accuracy<br>Finetuned on Public<br>Tested on Public | **0.9584**<br>[0.9564,<br>0.9607] | **0.9507**<br>[0.9477,<br>0.9531] | **0.948**<br>[0.9449,<br>0.9498] | **0.9463**<br>[0.9429,<br>0.9492] | **0.76** | **0.9934**<br>[0.993, 0.9938] | **0.9899**<br>[0.9892,<br>0.9904] | **0.9934**<br>[0.9859,<br>0.9875] | **0.9903**<br>[0.9895,<br>0.9908] | **0.6978** |
| Precision<br>Finetuned on Public<br>Tested on<br>SKMCH&RC | **0.5278**<br>[0.5142,<br>0.5495] | **0.6575**<br>[0.6323,<br>0.6746] | **0.6307**<br>[0.6051, 0.646] | **0.6442**<br>[0.6214,<br>0.6594] | **0.9214** | **0.7468**<br>[0.728, 0.7667] | **0.7368**<br>[0.7187,<br>0.7542] | **0.7468**<br>[0.728,<br>0.7667] | **0.74**<br>[0.7206,<br>0.7616] | **0.9156** |
| Recall<br>Finetuned on Public<br>Tested on<br>SKMCH&RC | **0.644**<br>[0.6195,<br>0.6726] | **0.7736**<br>[0.7538,<br>0.7906] | **0.7522**<br>[0.7321,<br>0.7664] | **0.7603**<br>[0.7413,<br>0.7745] | **0.9118** | **0.8214**<br>[0.81, 0.832] | **0.8278**<br>[0.8181,<br>0.8365] | **0.8214**<br>[0.81, 0.832] | **0.7941**<br>[0.7822,<br>0.8093] | **0.8716** |
| **F1**<br>**Finetuned on Public**<br>**Tested on**<br>**SKMCH&RC** | **0.5801**<br>[0.5629,<br>0.6019] | **0.7108**<br>[0.6923,<br>0.728] | **0.6861**<br>[0.6642,<br>0.6999] | **0.6974**<br>[0.6808,<br>0.7103] | **0.9097** | **0.7823**<br>[0.7729, 0.7961] | **0.7797**<br>[0.7691,<br>0.7913] | **0.7823**<br>[0.7729,<br>0.7961] | **0.766**<br>[0.7545,<br>0.7847] | **0.8853** |
| **Accuracy**<br>**Finetuned on Public**<br>**Tested on**<br>**SKMCH&RC** | **0.8906**<br>[0.8833,<br>0.8973] | **0.8705**<br>[0.8615,<br>0.8787] | **0.8498**<br>[0.8406,<br>0.8573] | **0.8502**<br>[0.8428,<br>0.8582] | **0.9118** | **0.9556**<br>[0.9511, 0.9597] | **0.9446**<br>[0.9391,<br>0.9497] | **0.9556**<br>[0.9511,<br>0.9597] | **0.9229**<br>[0.915,<br>0.9303] | **0.8716** |
| **Precision**<br>**Finetuned on**<br>**SKMCH&RC Tested**<br>**on SKMCH&RC** | **0.7826**<br>[0.7608,<br>0.8027] | **0.8328**<br>[0.8175,<br>0.8529] | **0.8098**<br>[0.7906,<br>0.8303] | **0.8243**<br>[0.8016,<br>0.8424] |  | **0.8449**<br>[0.8303, 0.8604] | **0.9115**<br>[0.8967,<br>0.9225] | **0.8987**<br>[0.8844,<br>0.9089] | **0.9093**<br>[0.8947,<br>0.9184] |  |
| **Recall**<br>**Finetuned on**<br>**SKMCH&RC Tested**<br>**on SKMCH&RC** | **0.7812**<br>[0.7639,<br>0.8007] | **0.8341**<br>[0.8208,<br>0.8516] | **0.8188**<br>[0.7994,<br>0.8388] | **0.8395**<br>[0.8252,<br>0.8543] |  | **0.8704**<br>[0.8556, 0.8846] | **0.9178**<br>[0.9048,<br>0.9314] | **0.9076**<br>[0.8981,<br>0.9214] | **0.9131**<br>[0.8967,<br>0.9275] |  |
| **F1**<br>**Finetuned on**<br>**SKMCH&RC Tested**<br>**on SKMCH&RC** | **0.7819**<br>[0.7659,<br>0.8017] | **0.8334**<br>[0.8204,<br>0.8521] | **0.8142**<br>[0.7993,<br>0.8343] | **0.8318**<br>[0.8136,<br>0.8468] |  | **0.8575**<br>[0.8449, 0.8717] | **0.9146**<br>[0.9015,<br>0.9263] | **0.9031**<br>[0.8912,<br>0.914] | **0.9112**<br>[0.8967,<br>0.9222] |  |
| **Accuracy**<br>**Finetuned on**<br>**SKMCH&RC Tested**<br>**on SKMCH&RC** | **0.9531**<br>[0.9502,<br>0.9571] | **0.9399**<br>[0.935,<br>0.9456] | **0.934**<br>[0.9281,<br>0.9407] | **0.9382**<br>[0.9319,<br>0.9431] |  | **0.9733**<br>[0.9697, 0.9769] | **0.9761**<br>[0.9733,<br>0.9792] | **0.9744**<br>[0.9720,<br>0.9774] | **0.9737**<br>[0.9700,<br>0.977] |  |
| **Precision**<br>**Finetuned on**<br>**SKMCH&RC Tested**<br>**on Public** | **0.4398**<br>[0.4315,<br>0.4523] | **0.6317**<br>[0.6217,<br>0.6429] | **0.6525**<br>[0.6432,<br>0.6632] | **0.6274**<br>[0.6172,<br>0.6384] |  | **0.6757**<br>[0.6654, 0.686] | **0.7317**<br>[0.7244,<br>0.7399] | **0.712**<br>[0.703,<br>0.7229] | **0.7302**<br>[0.724,<br>0.7378] |  |
| **Recall**<br>**Finetuned on**<br>**SKMCH&RC Tested**<br>**on Public** | **0.4068**<br>[0.3963,<br>0.4194] | **0.5321**<br>[0.5191,<br>0.5459] | **0.5948**<br>[0.5811,<br>0.6094] | **0.5623**<br>[0.5451,<br>0.5773] |  | **0.6291**<br>[0.6166, 0.6463] | **0.6417**<br>[0.6323,<br>0.6492] | **0.7112**<br>[0.7002,<br>0.7253] | **0.6335**<br>[0.6227,<br>0.646] |  |
| **F1**<br>**Finetuned on**<br>**SKMCH&RC Tested**<br>**on Public** | **0.4226**<br>[0.4146,<br>0.4337] | **0.5776**<br>[0.5671,<br>0.5879] | **0.6223**<br>[0.6121,<br>0.6336] | **0.593**<br>[0.5799,<br>0.6039] |  | **0.6515**<br>[0.643, 0.6625] | **0.6837**<br>[0.6774,<br>0.6899] | **0.7116**<br>[0.7043,<br>0.7204] | **0.6784**<br>[0.6722,<br>0.6871] |  |
| **Accuracy**<br>**Finetuned on**<br>**SKMCH&RC Tested**<br>**on Public** | **0.8199**<br>[0.8161,<br>0.8243] | **0.8269**<br>[0.8209,<br>0.832] | **0.8379**<br>[0.8315,<br>0.8421] | **0.8363**<br>[0.83,<br>0.842] |  | **0.9524**<br>[0.9503, 0.9541] | **0.9363**<br>[0.9337,<br>0.9386] | **0.9488**<br>[0.947,<br>0.9505] | **0.9343**<br>[0.9322,<br>0.9364] |  |

**Table 4:**
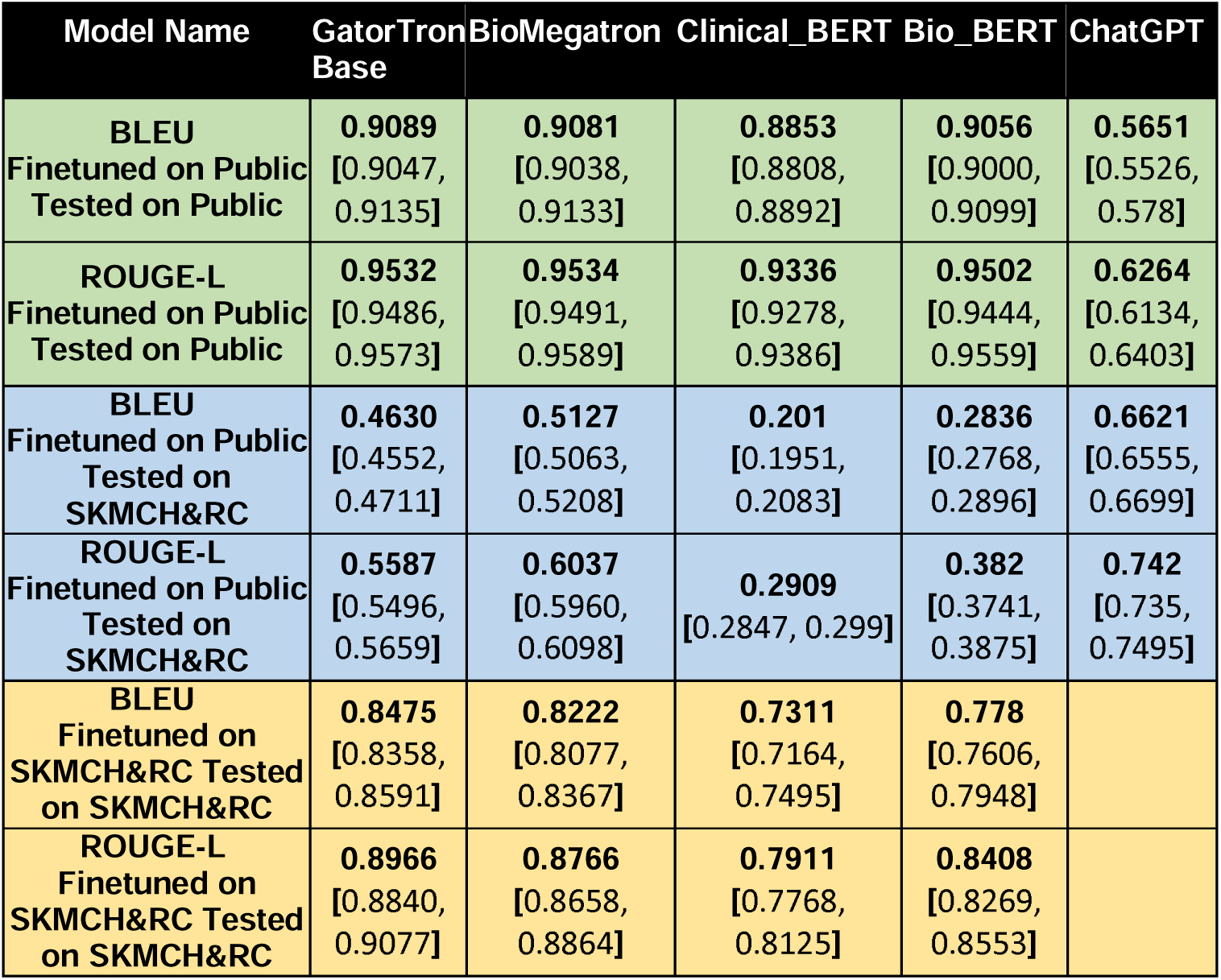
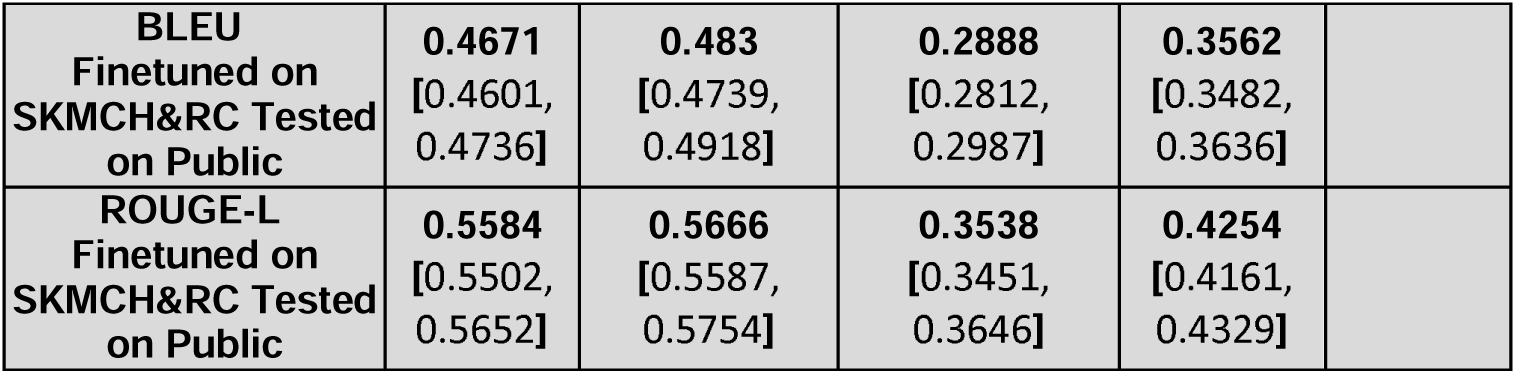
Question answering model performance for internal and external validation of LLMs fine-tuned on publicly available and SKMCH&RC datasets. In this table, each row indicates whether fine-tuning and testing were conducted on public or SKMCH&RC datasets.

In general, for open-source LLMs, including GatorTron, BioMegatron, ClinicalBERT, and BioBERT, when we fine-tuned the models on public data, their performance significantly reduced when tested on SKMCH&RC, and vice versa in both NLP tasks. This observation indicates the presence of bias in these models, likely stemming from inherent biases in the data sources on which these models were trained. Interestingly, ChatGPT, whose source training datasets are not fully disclosed, exhibited higher performance on SKMCH&RC compared to publicly accessible datasets. This pattern persisted across other performance metrics such as accuracy, precision, and recall for concept extraction and BLEU and ROUGE metrics for question answering task (Figure 2 a and b).

**Figure 2:** Difference between performance on public and SKMCH&RC data compared between models. a) compares 3-label performance, b) compares 9-label performance.

In general, LLMs fine-tuned on the SKMCH&RC training dataset resulted in the highest accuracy, precision, recall, and F1 score when tested on the SKMCH&RC test set. Specifically, the highest and lowest accuracy of 0.9531 (0.9502, 0.9571) and 0.934 (0.9281, 0.9407) belongs to GatorTron and ClinicalBERT respectively and the highest and lowest F1 score of 0.8334 (0.8204, 0.8521) and 0.7819 (0.7659, 0.8017) belongs to BioMegaTron and GatorTron in the dataset with 3-labels (I2B2). For the dataset with 9 labels (N2C2), BioMegaTron had the best performance with accuracy and F1 score of 0.9761 (0.9733, 0.9792), 0.9146 (0.9015, 0.9263) respectively, and GatorTron had the worst performance with 0.9733 (0.9697, 0.9769) and 0.8575 (0.8449, 0.8717).

For question answering task, GatorTron performs the best with BLEU and ROUGE-L scores of 0.8475 (0.8358, 0.8591) and 0.8966 (0.884, 9077) and ClinicalBERT was the worst performing with 0.7311(0.7164, 7495) and 0.7911 (0.7768, 0.8125). ChatGPT produced BLEU of 0.6621 (0.6555, 0.6699) and ROUGE-L of 0.742 (0.735, 0.7495)

Models fine-tuned on public datasets I2B2 and N2C2 resulted in the highest accuracy, precision, recall, and F1 score when internally validated on their respective test sets. In the I2B2 dataset, GatorTron demonstrated superior performance in terms of accuracy, achieving a score of 0.9584 (0.9564, 0.9607). ClinicalBERT outperformed other models in recall and F1 score, attaining scores of 0.8791 (0.8679, 0.888) and 0.8701 (0.8623, 0.8746), respectively. BioBERT exhibited the highest precision, recording a value of 0.0.8641 (0.8584, 0.8704). Conversely, ChatGPT displayed the lowest performance across all metrics, yielding scores of 0.76 for accuracy, 0.8114 for precision, 0.7667 for F1 score, and 0.76 for recall. For the N2C2 dataset, GatorTron emerged as the top performer in terms of accuracy, precision, F1 score with 0.9934 (0.993, 0.9938), 0.9555 (0.9516, 0.9599), and 0.959 (0.9556, 0.9628) respectively and BioBERT had the best Recall of 0.9678 (0.9653, 0.9714). In contrast, ChatGPT exhibited the least favourable performance on this dataset, achieving scores of 06978 for accuracy, 0.0.9124 for precision, 0.7496 for F1 score, and 0.6978 for recall.

Highest performing models for question answering in open dataset was GatorTron with the highest BLEU of 0.9089 (0.9047, 0.9135) and BioMegaTron with the highest ROUGE-L of 0.9534 (0.9491, 0.9589) and ClinicalBERT had the worst BLEU and ROUGE-L of 0.8853 (0.8808, 0.8892), and 0.9336 (0.9278, 0.9386) respectively. ChatGPT was evaluated with BLEU of 0.5651 (0.5526, 0.578) and ROUGE-L of 0.6264 (0.6134, 0.6403)

When models fine-tuned on public datasets I2B2 and N2C2 were evaluated on the SKMCH&RC test set with three labels, a significant decline in performance was observed compared to models fine-tuned specifically on SKMCH&RC, as illustrated in Figure 2a. This decline in performance persisted when evaluating the SKMCH&RC test set with nine labels (Figure 2b). A similar pattern was evident in the question answering task (Figure 2c).

Figure 3 presents the F1 scores of Language Models (LLMs) under different settings. Each LLM is depicted with a distinct colour, and the size of each circle corresponds to the F1 score of the models. The circles are divided by a “/”, where the text before the “/” indicates the dataset used for fine-tuning, and the text after the “/” signifies the datasets used for testing. Upon examination of the figure, a notable trend emerges: for each model designed to accommodate any number of labels, circles labelled with the same dataset for both fine-tuning and testing exhibit larger relative radii. This observed difference in radii can serve as an indicator of the dissimilarity in distributions between the two datasets, providing valuable insights into the impact of dataset variations on model performance.

**Figure 3:** Comparative Analysis of LLMs F1 Scores Across Varied Settings, Highlighting Dataset-Dependent Performance Disparities. The circle radius corresponds with F1 score – larger circles depict higher scores.

Figure 4 shows the label-specific misclassification performance for GatorTron, which had the largest drop in performance in internal (tested on I2B2) vs external validation (tested on SKMCH&RC). The left of Figure 4a showcases the absolute difference in model performances normalized by true labels, while the right highlights the differences between normalized confusion matrices based on predicted labels. Examining the diagonal elements of the matrices suggests differences in misclassification of all labels in the two testing datasets, with smaller differences observed in “B-Treatments.” In the 9-label dataset (Figure 4b), the smallest differences were in the classification of the “B-Drug”, “B-Form”, “B-Frequency”, “I-Duration”, and “I-Frequency” labels. A similar pattern was observed for the other LLMs.

**Figure 4:** Absolute Differences in Normalized Confusion Matrices between true and predicted labels. The left tables show normalized to true labels, and the right tables show normalized to predicted labels; a) shows the 3-label I2B2 dataset difference to SKMCH&RC data, b) shows the 9-label N2C2 and SKMCH&RC data.

Figure 5 displays two excerpts from sample notes within the SKMCH&RC dataset, each containing 9 labels (19 BIO labels), with true labels annotated by clinical experts and estimated labels generated by the GatorTron LLM. The 5a represents a snippet of text for which the LLM performed well in correctly predicting labels for the tokens, while 5b illustrates a case where the LLM performed poorly.

**Figure 5:** LLM labelling of 2 different clinical notes examples, demonstrating where the model performs well (a) and performs badly (b). Red boxes indicate misclassified labels; both examples were labelled by the GatorTron LLM using the 9-label classifications.

## Discussion

In this study, we tested openly available medical LLMs in a real-world clinical setting in South Asia. Internal and external validation of the performance of these LLMs was performed, with and without fine-tuning to the local EHR dataset. We further tested how well LLMs fine-tuned on the local dataset perform when tested on open-source medical datasets.

In general, models fine-tuned on open datasets performed poorly on the local EHR database. However, the same models, when fine-tuned on the local EHR database, performed well when tested on the local EHR database, albeit poorly on open datasets. Interestingly, ChatGPT, whose source training datasets are not fully disclosed (at the time of writing), was the exception to this trend, exhibiting better performance in terms of accuracy, F1 score, precision, recall, BLEU, ROUGE-L when tested on SKMCH&RC compared to publicly accessible datasets.

Our findings indicate that off-the-shelf LLMs can be biased when directly applied in real-world clinical settings. This is unsurprising given that the LLMs used in the study were trained on open-source datasets that are not representative of data from the real-world clinical setting we investigated. However, our results also showed it is possible to reduce such data-driven bias by fine-tuning the models on local data, under expert supervision. By doing so, the models can be made more equitably tailored to the local clinical needs.

The equitable use of LLMs entails several considerations. LLMs that do not perpetuate data-generated and algorithmic biases can make this powerful technology beneficial for diverse patient populations in resource-limited, overburdened settings. However, for these models to be applicable in a localised setting, the training data must be representative of these populations. We know that most research-ready medical data perpetuate the digital divide, overrepresenting high-income geographies and populations, which results in algorithmic assumptions about gender, race, geography and socio-economic status, etc. On the other hand, local data used in isolation is often sparse and ungeneralisable. As demonstrated in this study, fine-tuning is one way to reduce data-driven biases which can result in health inequity. It can also help to save time and resources over training an LLM from scratch, which can otherwise be prohibitively time and resource-intensive and requires expensive computing infrastructure which renders it inaccessible to healthcare providers in resource-limited settings. Tailoring to local data and context can also help alleviate the issue of hallucination and confabulation in LLMs, where a model can produce made-up or clinically nonsensical results. Reducing the likelihood of these outputs increases the validity and applicability of LLMs in health care and makes the models more trustworthy for clinicians.

However, there are several key challenges in making models equitable in practice. Real-world data such as EHR databases are not pre-designed for LLM applications. The manual labelling of data, and re-structuring of clinical notes to conform to the required formatting of LLMs is time-consuming and demanding for busy clinician experts, preventing rapid development of suitably accurate and unbiased models. Their production also requires close collaboration between clinicians, data scientists and IT specialists, which further complicates the generation of models. For example, it is particularly important to ensure that LLM-generated responses are rigorously validated and that any potential biases or inaccuracies are identified, communicated, and corrected with adequate transparency. In addition, ethical concerns around the sharing of patient data necessitate privacy-preserving measures for these data sources.

In this work, we fine-tuned locally running, privacy-preserving LLMs within a federated learning framework that precluded the need for patient data sharing. This is just one of the strengths of this research; to our knowledge, this is one of the few studies investigating the use of LLM technology in the global South, and the first study from Pakistan. All LLMs used in this work are publicly available (except for ChatGPT, which requires a paid subscription), rendering this approach reproducible in other settings without access and cost constraints. Models were tested on two forms of clinical notes (DS and SOAP notes) for the task of parsing clinical notes, making the approach disease-agnostic and generalisable. This work can be applied to further downstream tasks such as summarising and extracting key information for busy clinicians, answering medical questions, organising patient workups and other instances of clinical task-shifting.

Fair AI fundamentally relies on locally-driven co-creation of models that are 1) trained on locally representative bias-aware datasets, 2) account for algorithmic bias in the modelling process, 3) incorporate transparent reporting, and 4) are subjected to independent, context-aware internal and external validation to ensure local use and generalisability. The strengths of this study lie in its demonstration of how these models can be produced, presenting the feasibility and value of producing localised LLMs in this way.

Inevitably our study has some limitations. The findings reflect the performance of models on a random subset of clinical notes extracted from one EHR dataset in South Asia. While this EHR dataset has national coverage and is therefore representative of data from the general population of Pakistan in terms of socio-demographics and medical history, it represents a smaller-scale basis for LLM training. The key to successful health innovation lies in its reliability and perception as fit for use by the populations of interest and key stakeholders. Alongside increasing the scale of training data, future work should also focus on the challenges of creating clinical LLMs trusted by doctors, nurses, and front-line staff, and how these users envision the future role of these models in patient care. The testing of these models in patient care will not only allow further analysis of their technical validity but also allow patients and clinicians to discuss the cultural and ethical acceptability of novel health-focused LLM technology in local health settings. This is an important first step to ensure that local stakeholders have agency and ownership in the development of transformative health technology, including LLMs, going forwards, in ways that make them safe, fair and beneficial for all.

## Conclusions

Despite the elevated interest in LLMs, their assessment in real-world clinical settings is lacking. This study evaluates the feasibility of equitable access and use of LLMs for clinical decision-making by assessing the performance of medical LLMs on a local dataset from a hospital in Pakistan. Given that most medical datasets suffer from the digital divide, we tested and independently validated LLMs on a large local EHR database. The database was first labelled by a team of clinical experts from the setting in context through expert consensus to review to contest and redress algorithmic decisions. To minimise algorithmic bias, we compared model performance with and without fine-tuning the LLMs to the local dataset.

We recognise that equitable use of health innovations extends well beyond considerations of technological bias; it necessitates consideration of clinical explainability, acceptability, trust and local ownership such as can be assessed through regular stakeholder engagement and interaction. This is an important avenue for future research to ensure clinical practitioners, decision-makers, and patients and carers have agency and ownership in the development and implementation of transformative health technology including LLMs going forward.

In conclusion, our findings highlight the presence of data-driven and algorithmic biases in existing publicly available clinical LLMs predominantly pre-trained on data from global north settings. This work indicates that pre-trained LLMs can be tailored for parsing clinical notes in specific clinical contexts, but only through careful consideration of multi-dimensional biases. It emphasizes the need for continued scrutiny of LLMs and suitable corrective measures, such as regular fine-tuning on local data under the supervision of local context-aware clinical experts.

## Data Availability

Aggregate data will be made freely available on study website. No patient-level data sharing is permitted as per ethics approval.

## Acknowledgements

This study was funded by Bill & Melinda Gates Foundation (INV-062576). We would like to acknowledge Amelia M. Doran from the University of Oxford for editing support.

